# Human phospho-signaling networks of SARS-CoV-2 infection are rewired by population genetic variants

**DOI:** 10.1101/2021.11.22.21266712

**Authors:** Diogo Pellegrina, Alexander T. Bahcheli, Michal Krassowski, Jüri Reimand

## Abstract

SARS-CoV-2 infection hijacks signaling pathways and induces protein-protein interactions between human and viral proteins. Human genetic variation may impact SARS-CoV-2 infection and COVID-19 pathology; however, the role of genetic variation in these signaling networks remains uncharacterized. We studied human single nucleotide variants (SNVs) affecting phosphorylation sites modulated by SARS-CoV-2 infection, using machine learning to identify amino acid changes altering kinase-bound sequence motifs. We found 2033 infrequent phosphorylation-associated SNVs (pSNVs) that are enriched in sequence motif alterations, potentially reflecting the evolution of signaling networks regulating host defenses. Proteins with pSNVs are involved in viral life cycle processes and host responses, including regulators of RNA splicing and interferon response, as well as glucose homeostasis pathways with potential associations with COVID-19 co-morbidities. Certain pSNVs disrupt CDK and MAPK substrate motifs and replace these with motifs recognized by Tank Binding Kinase 1 (TBK1) involved in innate immune responses, indicating consistent rewiring of infection signaling networks. Our analysis highlights potential genetic factors contributing to the variation of SARS-CoV-2 infection and COVID-19 and suggests leads for mechanistic and translational studies.

## INTRODUCTION

Severe acute respiratory syndrome coronavirus 2 (SARS-CoV-2) and the coronavirus disease 2019 (COVID-19) pandemic has caused millions of deaths worldwide and continues to evolve as more pathogenic and infectious variants of the virus emerge. The clinical manifestations and outcomes of COVID-19 are complex, ranging from asymptomatic infection to fatal respiratory and multi-organ failure, as well as long-term symptoms after recovery. Risk factors of severe disease include advanced age, a weakened immune system, and pre-existing health conditions such as hypertension, diabetes, and obesity ^1^. Ethnic and demographic patient characteristics that are at least partially explained by socio-economic factors also affect disease risk ^2-4^. Recent genome-wide association studies (GWAS) of COVID-19 have shed light on human genetic variation contributing to SARS-CoV-2 disease burden and mortality rates, highlighting genes associated with ABO blood groups, antiviral pathways and tyrosine kinase signaling ^5-7^. However, GWAS findings often occur in intergenic regions where molecular mechanisms remain elusive, and rare variants potentially contributing to disease are challenging to detect. Thus, additional work is needed to find and interpret genetic variants contributing to SARS-CoV-2 infection and COVID-19 pathology.

Molecular interaction networks are perturbed by host-pathogen interactions and host disease mutations that either disable proteins in the networks or alter their interactions ^8^. Phosphorylation is a key component of cellular signaling networks that acts as a reversible molecular switch controlling protein function and interactions. This post-translational modification (PTM) is conducted by kinases that recognize sequence motifs at protein phosphorylation sites (*i*.*e*., phosphosites). SARS-CoV-2 infection alters phosphorylation networks in host cells by promoting casein kinase 2 (CK2) and mitogen-activated protein kinase (MAPK) pathways and inhibiting mitotic kinases, resulting in cell cycle arrest and cytoskeletal changes to favor virus pathology ^9^. Phosphorylation networks also control host anti-viral and immune responses to SARS-CoV-2 infection. For example, the TANK Binding Kinase 1 (TBK1) and IKK-epsilon kinase of the NF-κB pathway initiate innate antiviral response by phosphorylating interferon regulatory factors (IRF) that regulate interferon genes ^10,11^. Interferons activate JAK-STAT, p38 MAPK, and PI3K/Akt signaling pathways that are dysregulated in SARS-CoV-2 infection and COVID-19 ^9,12,13^. SARS-CoV-2 proteins bind TBK1 to suppress host immune responses ^14,15^.

NF-κB hyperactivation has been associated with cytokine storms in COVID-19 where excessive production of proinflammatory cytokines can cause fatal damage to the host ^16^. Besides host protein, post-translational modifications of virus proteins may also contribute to the complexity of infection and host anti-viral responses ^17^.

Genetic variants involved in human disease are known to affect kinase signaling networks. For instance, inherited disease mutations and somatic mutations in cancer driver genes frequently erase phosphosites or alter kinase binding motifs, potentially causing rewiring of kinase signaling ^18-21^. Conversely, phosphosites tend to have lower genetic variation in the human population, underlining the importance of conserved phosphosites in evolution ^18,22^. This suggests that human genetic variation of signaling networks responding to the novel SARS-CoV-2 infection may contribute to a range of symptoms, disease severity, and long-term outcomes of COVID-19 patients.

We hypothesized that genetic variation in phosphosites differentially phosphorylated in SARS-CoV-2 infection can alter kinase signaling interactions and thereby reveal mechanistic insights into the genes and pathways involved in infection and disease. We studied the gnomAD dataset ^23^, the largest uniformly processed map of human genetic variation comprising exome sequencing data of more than 100,000 individuals, and mapped missense single nucleotide variants (SNVs) to human protein phosphosites responding to SARS-CoV-2 infection ^9^. With a machine learning approach, we uncovered hundreds of phosphorylation-associated SNVs (pSNVs) that modify kinase-bound sequence motifs and potentially rewire kinase-substrate interactions. Our study helps decipher the role of human genome variation in virus responses and disease outcomes and enables advances in therapy and biomarker development.

## RESULTS

### SARS-CoV-2 associated phosphosites are enriched in pSNVs rewiring kinase binding motifs

To examine the human genetic variation in signaling networks responding to SARS-CoV-2 infection, we studied exome sequencing data of 124,748 individuals from 16 populations in the gnomAD dataset ^23^, focusing on 1,111,194 amino acid substitutions that were observed at least once per 10,000 individuals in at least one of the populations (AF_popmax_ > 10^−4^). We defined phosphorylation-associated SNVs (pSNVs) as missense SNVs (*i*.*e*., amino acid substitutions) that affected 1530 host phosphosites differentially phosphorylated in SARS-CoV-2 infection based on an earlier phosphoproteomics study (*FDR* < 0.05; 24h timepoint) ^9^. We mapped pSNVs in flanking windows of ±7 amino acid residues around phosphorylated residues to study sequence motifs bound by kinases (**Figure 1A**). To predict the functional impact of pSNVs on protein phosphorylation, we evaluated the effects of pSNVs on 125 types of sequence motifs bound by kinases of 77 families. We used MIMP ^24^, a machine learning method trained on kinase binding sequences, that evaluates whether a pSNV disrupts an existing motif or creates a new motif relative to the reference protein sequence. Higher Bayesian posterior probabilities computed by MIMP reflect an increased likelihood of pSNVs causing rewiring of sequence motifs.

**Figure 1.**
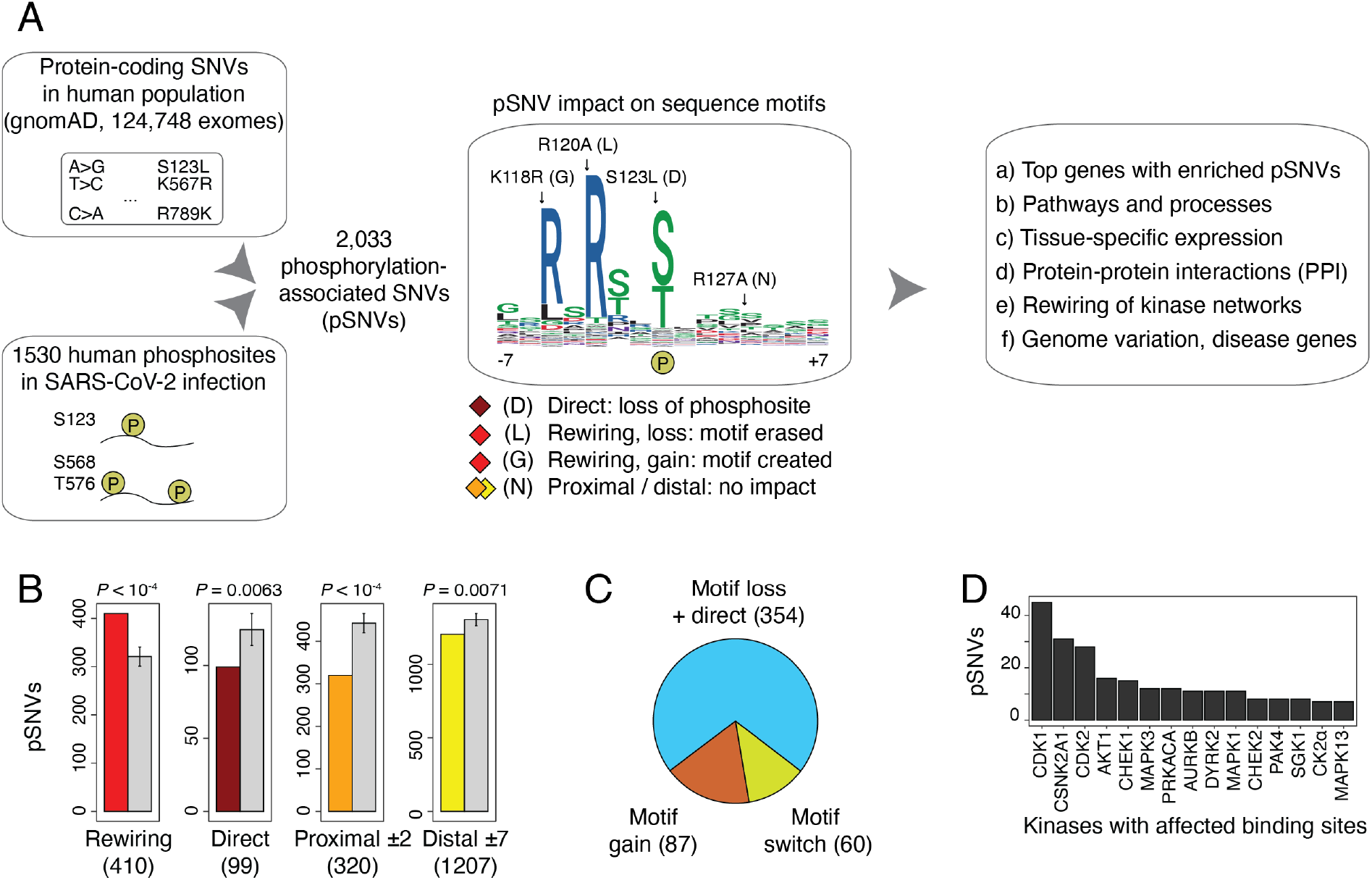
Phosphorylation-associated single nucleotide variants (pSNVs) in human phosphosites of SARS-CoV-2 infection. **A**. Overview of analysis. Missense SNVs in the human population (allele frequency AF_popmax_ ≥ 10^−4^ in gnomAD) were mapped to host protein phosphosites significantly differentially phosphorylated in SARS-CoV-2 infection ^9^. Sequence motif analysis was used to evaluate pSNV impact to predict gains and losses of kinase binding sites. Genes, pathways, and molecular interaction networks with frequent pSNVs were analyzed. **B**. pSNVs of four functional classes were studied: (1) motif-rewiring pSNVs altering sequence motifs in kinase binding sites (MIMP posterior *prob* > 0.5), (2) direct pSNVs altering phosphorylated residues, and (3-4) pSNVs with no motif-rewiring predictions proximal and distal to phosphosites (±1-2 and ±3-7 residues, respectively). Expected numbers of pSNVs (grey) represent control phosphosites sampled randomly from the proteome. Empirical P-values and pSNV counts are shown. pSNVs predicted to cause rewiring of sequence motifs are enriched compared to controls (shown in red). **C**. Impact of motif-rewiring pSNVs on protein phosphorylation. Pie chart shows pSNVs predicted to cause loss of phosphosites (*i*.*e*., pSNVs substituting phospho-residues and/or disrupting sequence motifs; blue), gain of phosphosites (*i*.*e*., pSNVs inducing new sequence motifs; orange), and motif switches (pSNVs erasing one motif and inducing another motif). **D**. A subset of pSNVs occur at high-confidence binding sites of kinases. Bar plot shows the top 15 kinases whose binding sites have the largest numbers of pSNVs.

We found 2,033 pSNVs in 987 SARS-CoV-2 -associated phosphosites and 693 genes and assigned these to four classes based on their predicted functional impact (**Figure 1B, Supplementary Table 1**). First, *direct* pSNVs (99 or 5%) replaced central phospho-residues and caused loss of phosphosites, likely representing the pSNV class of highest impact. Second, *motif-rewiring* pSNVs (410 or 20%) created or disrupted sequence motifs, potentially causing gains or losses kinase binding at the phosphosites (MIMP posterior *prob* > 0.5). The two remaining classes of pSNVs lacked functional predictions based on sequence analysis and included 1527 (75%) pSNVs annotated as *proximal* or *distal* to the closest phosphosite. When combining direct and motif-rewiring pSNVs, loss of phosphorylation through motif disruption or phospho-residue replacement was predicted for 354 pSNVs. 87 pSNVs induced new sequence motifs at SARS-CoV-2-associated phosphosites and were predicted to cause gain of phosphorylation (**Figure 1C, Supplementary Table 2**). Interestingly, 60 pSNVs caused motif switching where the pSNV simultaneously disrupted one motif and replaced it with another motif at the same phosphosite. Overall, hundreds of pSNVs in the human population are predicted to alter SARS-CoV-2 driven signaling in host cells through the range of kinase sequence motifs we studied.

We evaluated the statistical significance of these potentially functional pSNVs by randomly sampling human phosphosites as controls. Compared to experimentally determined phosphosites in the human proteome, SARS-CoV-2 associated phosphosites were enriched in motif-rewiring pSNVs (410 pSNVs observed *vs*. 321 ± 21 pSNVs expected; *P* < 10^−4^) (**Figure 1B**). In contrast, direct pSNVs replacing central phospho-residues, as well as proximal and distal pSNVs lacking motif-based predictions were found less frequently in SARS-CoV-2 associated phosphosites. Some pSNVs (213 or 10%) occurred at well-defined phosphosites where the phosphorylating kinases have been identified previously, such as cyclin dependent kinases, casein 2 kinases, checkpoint kinases, and MAP kinases (**Figure 1D**), in agreement with studies of the human signaling network modulated upon SARS-CoV-2 infection ^9^ and adding confidence to our sequence-based predictions of pSNV impact. Enrichment of motif-rewiring pSNVs suggests that the signaling network responding to SARS-CoV-2 infection in humans may vary structurally such that certain kinase-substrate interactions are gained or lost some individuals. This may reflect positive evolutionary selection in adaptation to viral infection and potentially cause variation in SARS-CoV-2 infection and COVID-19 disease course.

### Top genes with pSNVs are involved in RNA splicing, virus infection and host immune response

We studied the genes most affected by pSNVs in SARS-CoV-2-associated phosphosites by evaluating the null hypothesis that none of the pSNVs per gene caused sequence motif alterations in the resulting protein. We computed significance scores that assigned more weight to genes with multiple pSNVs affecting kinase binding motifs or replacing central phospho-residues, as log-sums of complementary posterior probabilities. Finally, the significance scores were adjusted for multiple testing using the false discovery rate (*FDR*) and filtered to select genes with significant pSNVs.

Gene prioritization revealed 77 genes with pSNVs predicted to alter protein phosphorylation through central residue or sequence motif alterations (*FDR* < 0.1), collectively including nearly one third of pSNVs (**Figure 2A, Supplementary Table 3**). RNA splicing and anti-viral defense responses were prominent among the functions of top genes. The two most significant genes, *SRRM2* and *BUD13*, encode subunits of the spliceosome complex and include 83 and 11 pSNVs, respectively (*FDR* < 10^−23^). SRRM2 is involved in human immunodeficiency virus (HIV) pathogenesis via alternative splicing ^25^. BUD13 regulates the antiviral transcription factor *IRF7* and interferon I response upon RNA-virus infection ^26,27^. Interferon signaling triggers the first response of host cells to viral infection and activates the immune system of adjacent cells to suppress viral replication. The host RNA splicing machinery is targeted by SARS-CoV-2 proteins to disrupt splicing and impair host gene translation ^28,29^. Another related candidate gene *BCLAF1* encodes a pro-apoptotic transcription and splicing factor and includes 18 pSNVs (*FDR* = 4.2 × 10^−6^). Apoptosis is triggered in infected host cells as a final form of cellular defense, while activation of anti-apoptotic pathways is a strategy to maximize viral replication ^30^. BCLAF1 is also involved in type I interferon signaling and regulates antiviral gene expression upon virus infection ^31^. The essential role of BCLAF1 in lung development ^32^ and expression in lung cells suggests its activity in airway tract tissues affected by SARS-CoV-2 infection. Thus, some top genes with pSNVs are involved in core host cellular processes of virus infection and host immune response.

**Figure 2.**
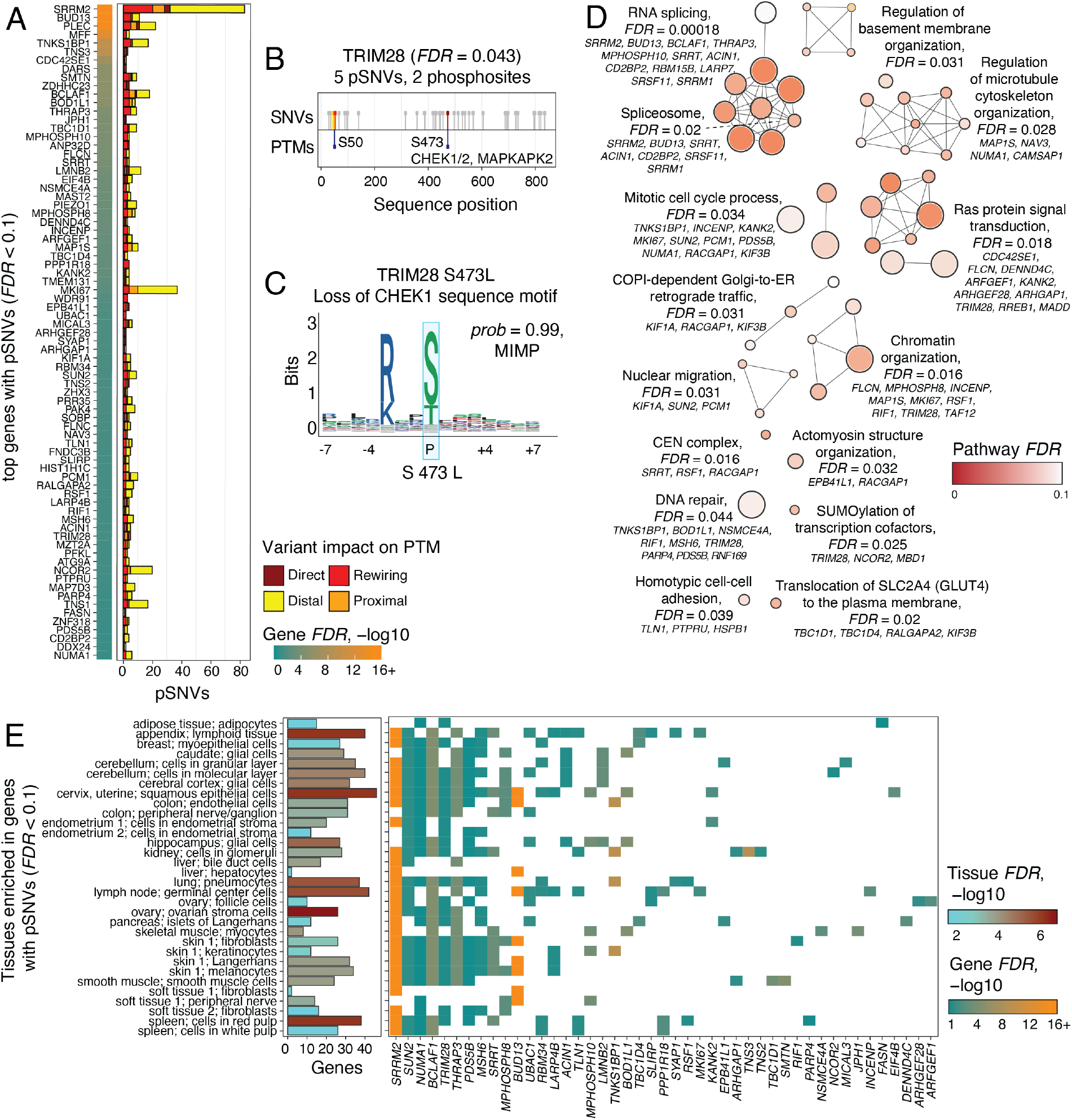
Top genes, pathways, and tissue-specific expression patterns of SARS-CoV-2 associated pSNVs. **A**. Top genes with pSNVs predicted to alter phosphosites modulated by SARS-CoV-2 infection (*FDR* < 0.1). Genes are scored by the probability of at least one pSNV altering SARS-CoV-2-associated phosphosites. **B**. Example of a top gene with pSNVs: TRIM28, a transcriptional and epigenetic regulator involved in innate immune response, includes five pSNVs in two phosphosites: S473 phosphorylated by CHEK and MAPKAP2 kinases, and the phosphosite S50 with no known kinase. **C**. The pSNV S473L in TRIM28 substitutes the phospho-residue and disrupts a CHEK1 sequence motif with a high-confidence prediction of motif alteration (MIMP posterior *prob* > 0.99). the pSNV potentially alters the regulation of interferons by TRIM28. **D**. pSNVs are enriched in biological processes and molecular pathways (ActivePathways, *FDR* < 0.1). The enrichment map is a network of pathways and processes as nodes that are connected by edges if the pathways share many genes. Each subnetwork represents a distinct functional theme. Top genes with pSNVs are listed for each theme and include additional genes found in the pathway analysis (gene *FDR* < 0.25). **E**. Tissue-specific expression of genes with frequent pSNVs, as detected by enrichment analysis of Human Protein Atlas gene sets (ActivePathways, *FDR* < 0.1). Bar plot (left) shows all the genes with pSNVs expressed in each tissue. Grid plot (right) shows the tissues where the top genes with pSNVs are expressed (gene *FDR* < 0.1).

We highlighted the transcriptional repressor *TRIM28* with five pSNVs as a gene of interest (*FDR* = 0.042) (**Figure 2B-C**). TRIM28 increases interferon beta and pro-inflammatory cytokine production in response to avian virus infection in lung epithelial cells through phosphorylation of the amino acid residue S473 ^33^. This phosphosite is also modified upon SARS-CoV-2 infection in Vero6 cells ^9^. One direct pSNV in TRIM28 affects the S473 phosphosite: the amino acid substitution S473L removes the central phospho-residue and causes loss of phosphorylation. The site S473 is phosphorylated by checkpoint kinases (CHEK1/2) and the MAPKAP2 kinase in DNA damage response and interferon activation ^33,34^. In agreement with those experimentally verified kinase binding sites, MIMP analysis predicts that the sequence motifs of the same kinases are disrupted by the pSNV, including motifs of CHEK1, CHEK2 and MAPKAP2 (MIMP posterior *prob* ≥ 0.99) (**Figure 2C**). Another pSNV in TRIM28, R472C, occurs upstream of the S473 phosphosite; however, unlike S473, this substitution does not cause significant motif alterations. A recent study showed that knockdown of *TRIM28* activates ACE2 and leads to increased host cell entry of SARS-CoV-2 ^35^. TRIM28 expression also correlates with interferon levels, being lower in children with severe COVID-19 disease and multisystem inflammatory syndrome (MIS-C) compared to uninfected children and those with mild disease ^36^. The pSNVs in the phosphosites of TRIM28 may alter the signaling of this protein and suppress the activation of immune response. Further study of this protein and its pSNVs may offer mechanistic insights to its role in SARS-CoV-2 infection and COVID-19.

### Genes with frequent pSNVs are broadly expressed and enriched in host processes of virus life cycle

To interpret the human genetic variation of signaling networks responding to SARS-CoV-2 infection, we asked if the genes with frequent pSNVs converged to biological processes, molecular pathways, and protein complexes. Pathway enrichment analysis highlighted 50 gene sets with frequent pSNVs and also captured additional genes with pSNVs that remained sub-significant in the gene-focused analysis (*FDR* < 0.1 from ActivePathways ^37^) (**Figure 2D, Supplementary Table 4**). Post-transcriptional regulation and RNA splicing represented the largest group of pathways and included the spliceosome complex with 120 pSNVs in seven proteins (CORUM:351, *FDR* = 0.020), extending our observations from the top gene list. Second, gene sets involved in basement membrane, cytoskeleton and microtubule organization were enriched, perhaps reflecting the structural changes in cellular organization upon virus infection ^9^. The third major group involved Ras and Rho GTPase signaling proteins that respond to extracellular stimuli to activate diverse cellular pathways such as proliferation, migration, apoptosis, and cell adhesion. The Reactome pathway *COPI-dependent Golgi-to-ER retrograde traffic* (*FDR* = 0.031; R-HSA-6811434) with 19 pSNVs in eight genes included the kinesins *KIF1A* and *KIF3B* involved in intracellular transport, and the GTPase signaling protein *RACGAP1*. SARS-CoV-2 uses the host translational machinery in the endoplasmic reticulum (ER) for replication, resulting in increased ER stress and activation of unfolded protein response ^30,38^. Gene sets related to chromatin organization, DNA repair, cell adhesion, and mitotic cell cycle processes were also enriched. Thus, pSNVs affect multifunctional proteins involved in core host processes of the virus life cycle.

Genes with frequent pSNVs genes were expressed in diverse human tissues and cell types according to functional enrichment analysis tissue-specific gene expression signatures in Human Protein Atlas ^39^ (ActivePathways *FDR* < 0.1) (**Figure 2E, Supplementary Table 4**). The prioritized genes were often expressed in lung pneumocytes and their squamous epithelial precursors, confirming their relevance to respiratory tissues affected by SARS-CoV-2 infection. The genes were also often expressed in lymph nodes and spleen, which are damaged by SARS-CoV-2 infection ^40^. Brain and nervous system tissues, including cerebral cortex, hippocampus, and peripheral nerves were also identified among enriched gene expression signatures, in line with evidence of broad cellular perturbations of brain tissues in severe COVID-19 ^41^. Gene expression signatures of skin, kidney, pancreas, colon, liver, and female reproductive tissues were also identified. Tissue-specific enrichments were partially driven by genes with high expression in most tissues, such as those involved in RNA splicing (*SRRM2, BCLAF1*), interferon regulation (*TRIM28*), cell cycle (*NUMA1, SUN2*) and DNA damage response (*MSH6*). SARS-CoV-2 affects diverse human tissues directly through the ACE2 receptor and indirectly through inflammation and immune response dysregulation, causing broad organ damage in severe disease ^42^. Pathway annotations and expression patterns of genes with pSNVs suggest these multifunctional genes and their genetic variants may contribute to SARS-CoV-2 infection and COVID-19 through various pathways and mechanisms.

### Protein-protein interaction networks with frequent SARS-CoV-2-associated pSNVs

We performed a network analysis of protein-protein interactions (PPI) to understand the functional context of the top genes with pSNVs (**Figure 3A, Supplementary Table 5**). We included 30 physical PPIs of the 77 top human proteins, and 139 PPIs of SARS-CoV-2 proteins interacting with the top human proteins collected in the BioGRID database ^43^. We also included 29 high-confidence kinase-substrate interactions with specific SARS-CoV-2-associated phosphosites, using data from the ActiveDriverDB database ^44^. Kinase-substrate interactions were limited to phosphosites with at least one pSNV. To evaluate the significance of this PPI network, we sampled subnetworks from the human proteome using random sets of phosphoproteins as controls.

**Figure 3.**
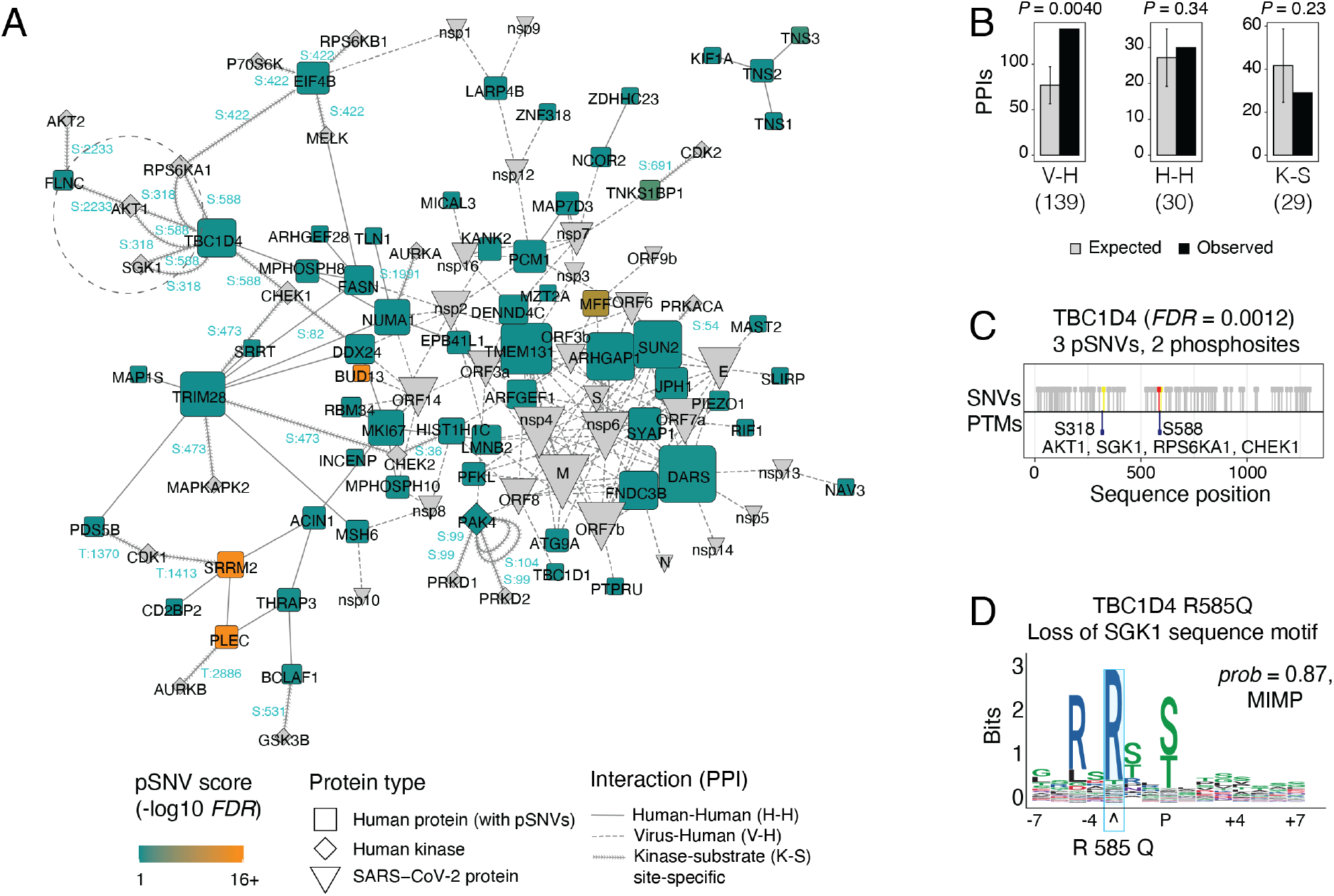
Protein-protein interactions (PPI) of genes with pSNVs in SARS-CoV-2 associated phosphosites. **A**. The PPI network includes three classes of proteins: (a) human proteins encoded by top genes with pSNVs (*FDR* < 0.1; blue and orange squares), (b) proteins encoded by the SARS-CoV-2 virus (grey wedges), and (c) human kinases (grey diamonds) known to bind SARS-CoV-2-associated phosphosites with pSNVs. Three types of interactions are shown: PPIs of the top human proteins (H-H; solid line), PPIs of the top human proteins and SARS-CoV-2 proteins (V-H; dashed line), and site-specific kinase-substrate phosphorylation events at sites with pSNVs (K-S; arrow line). The phosphosites with pSNVs bound by specific kinases are shown in blue (*e*.*g*., S99). Node size corresponds to the number PPIs per protein (*i*.*e*., node degree). **B**. Enrichment analysis of PPIs. The PPI network is significantly enriched in host-virus interactions of top human proteins and SARS-CoV-2 proteins (i.e., V-H interactions; left). Human-human PPIs (H-H) and site-specific kinase-substrate interactions (K-S) are not enriched. Expected PPI counts were derived from control protein sets drawn randomly from the human phospho-proteome. Empirical P-values are shown. **C**. Example of kinase-substrate interactions altered by pSNVs in phosphosites of SARS-CoV-2 infection (dotted circle in panel A). The GTPase signaling protein TBC1D4 involved in glucose homeostasis and transport, diabetes, and obesity, includes three pSNVs in the phosphosites S318 and S588 phosphorylated by the kinases AKT1, CHEK1, RPS6KA1 and SGK1. **D**. The pSNV R585Q in TBC1D4 substitutes R585 adjacent to the phospho-residue and disrupts the sequence motif of SGK1, potentially disrupting glucose homeostasis regulated by TBC1D4.

Human proteins with pSNVs interacted with viral proteins significantly more often than expected from the human phospho-proteome (139 observed PPI *vs*. 77 ± 20 expected (±1 s.d.); permutation *P* = 0.0040) (**Figure 3B**). The most frequently interacting human proteins (DARS, TMEM131, ARHGAP1, SUN2) and viral proteins (M, nsp4, nsp6, orf7a, orf7b) each had at least 10 interactions in the network. SUN2 with 9 pSNVs (*FDR* = 0.021) encodes a nuclear membrane protein whose over-expression blocks HIV-1 replication and induces changes in nuclear shape ^45^. RNA-binding proteins and spliceosome subunits such as BUD13, DDX24, and RBM34 interact with orf14, a currently uncharacterized SARS-CoV-2 protein ^26,46^. DDX24 is involved in RNA packaging of HIV into virions ^47^. RBM34 has been implicated in Middle Eastern Respiratory Syndrome (MERS) infections via interactions with the viral protein nsp3.2 ^48^. The SARS-CoV-2 non-structural proteins nsp4 and nsp6, frequently interacting with top pSNV-enriched host proteins, localize to ER, and are involved in autophagy and viral replication ^49,50^. nsp6 suppresses interferon regulatory factor 3 (IRF3) phosphorylation by binding TANK binding kinase 1 (TBK1) ^15^. Thus, the human proteins with frequent pSNVs are involved in infection and pathogenesis through interactions with viral proteins, suggesting that some pSNVs could impact SARS-CoV-2 virus-host protein interactions and signaling pathways.

We examined the site-specific kinase-substrate interactions for additional details of pSNVs. The enriched pathway *Translocation of SLC2A4 (GLUT4) to the plasma membrane* (Reactome: R-HSA-1445148) with pSNVs in *TBC1D1, TBC1D4, RALGAPA2, KIF3B* and others (*FDR* = 0.02; ActivePathways) identified in our pathway analysis is also apparent in the PPI network. *TBC1D4* (*i*.*e*., AS160), encodes a GTPase-activating protein that regulates glucose homeostasis through insulin-dependent trafficking of GLUT4 ^51^. Protein-coding SNVs in TBC1D4 and TBC1D1 have been associated with insulin resistance, obesity, and type II diabetes ^52,53^. Type II diabetes and obesity are SARS-CoV-2 comorbidities and may implicate certain individuals and populations as more susceptible to severe COVID-19 ^45^. In our data, TBC1D4 includes three pSNVs that affect the SARS-CoV-2-associated phosphosites S318 and S588 (*FDR* = 0.0011). The sites are phosphorylated by AKT1, SGK1, RPS6KA1, and CHEK1 kinases based on earlier experimental studies ^54-56^ (**Figure 3C**). The pSNV R585Q disrupts sequence motifs in the TBC1D4 phosphosite S588, including motifs bound by the same kinases (CHEK1, SGK1, RPS6KA1) (**Figure 3D**). TBC1D4 phosphorylation at S588 is known to regulate GLUT4 translocation and subsequent activity in adipocytes ^51^. Thus, the predicted motif disruption via pSNVs matches the kinases binding the phosphosite. The related protein TBC1D1 has nine pSNVs in the phosphosites S614, S627, and S660, including four pSNVs potentially disrupting phosphorylation (*FDR* = 3.8 × 10^−5^); however, the kinases binding these phosphosites are unknown. This example suggests that some pSNVs provide mechanistic hypotheses for studying SARS-CoV-2 comorbidities.

### Motif-switching pSNVs exchange CDK and MAPK kinase targets in favor of TBK1

To characterize the effect of pSNVs on rewiring SARS-CoV-2 phosphorylation networks, we studied the kinase families whose sequence motifs were most affected by pSNVs in top genes. We computed significance scores for kinase-substrate pairs by accounting for all high-confidence motif predictions, giving a higher priority to kinase-substrate pairs where the same type of sequence motif was affected by multiple pSNVs of the protein. This strategy included similar motifs of evolutionarily related kinase families and led to partially redundant scoring of kinase motifs. Collectively, we included 217 pSNVs with 3360 motif-rewiring predictions covering 64 kinase families.

Hierarchical clustering of motif-rewiring pSNVs revealed two major clusters of kinases and substrate proteins (**Figure 4A**). The first cluster was characterized by pSNVs causing losses of binding motifs of cyclin dependent kinases (CDKs) and mitogen associated kinases (MAPK; p38, Jnk, Erk) in genes encoding the proliferation marker MKI67, DNA damage response protein MSH6, spliceosome subunit SRRT, interferon regulator BCLAF1, the translation initiation factor EIF4B, and others. For instance, sequence binding motifs of CDK7 were lost through 56 pSNVs and gained through two pSNVs (**Figure 4B**). SARS-CoV-2 infection is known to promote activation of host MAPK p38 kinases and inhibition of CDKs ^9^. The second, larger cluster involved motif rewiring of Protein Kinase A, CAMK, checkpoint, and PI3K/Akt kinase. The PI3K/Akt pathway controls cell proliferation, survival, and suppression of apoptosis, and responds to extracellular signals of cytokines and growth factors via the mTOR pathways. Checkpoint kinases respond to DNA damage by blocking cell proliferation and inducing cell death. Among the top proteins, motifs bound by Protein Kinase A (PRKACA) and CAMK2A were affected by 98 pSNVs, most of which led to motif losses or replacements of central phospho-residues (**Figure 4B**). Motif-rewiring pSNVs in this cluster occur in transcriptional and post-transcriptional regulators such as PLEC, BUD13, THRAP3, and TRIM28, among others. The spliceosome subunit SRRM2, with the largest number of pSNVs in our analysis, included motif-rewiring pSNVs affecting kinases of both clusters. Therefore, pSNVs may reconfigure kinase signaling networks of defense processes and host components of the viral life cycle.

**Figure 4.**
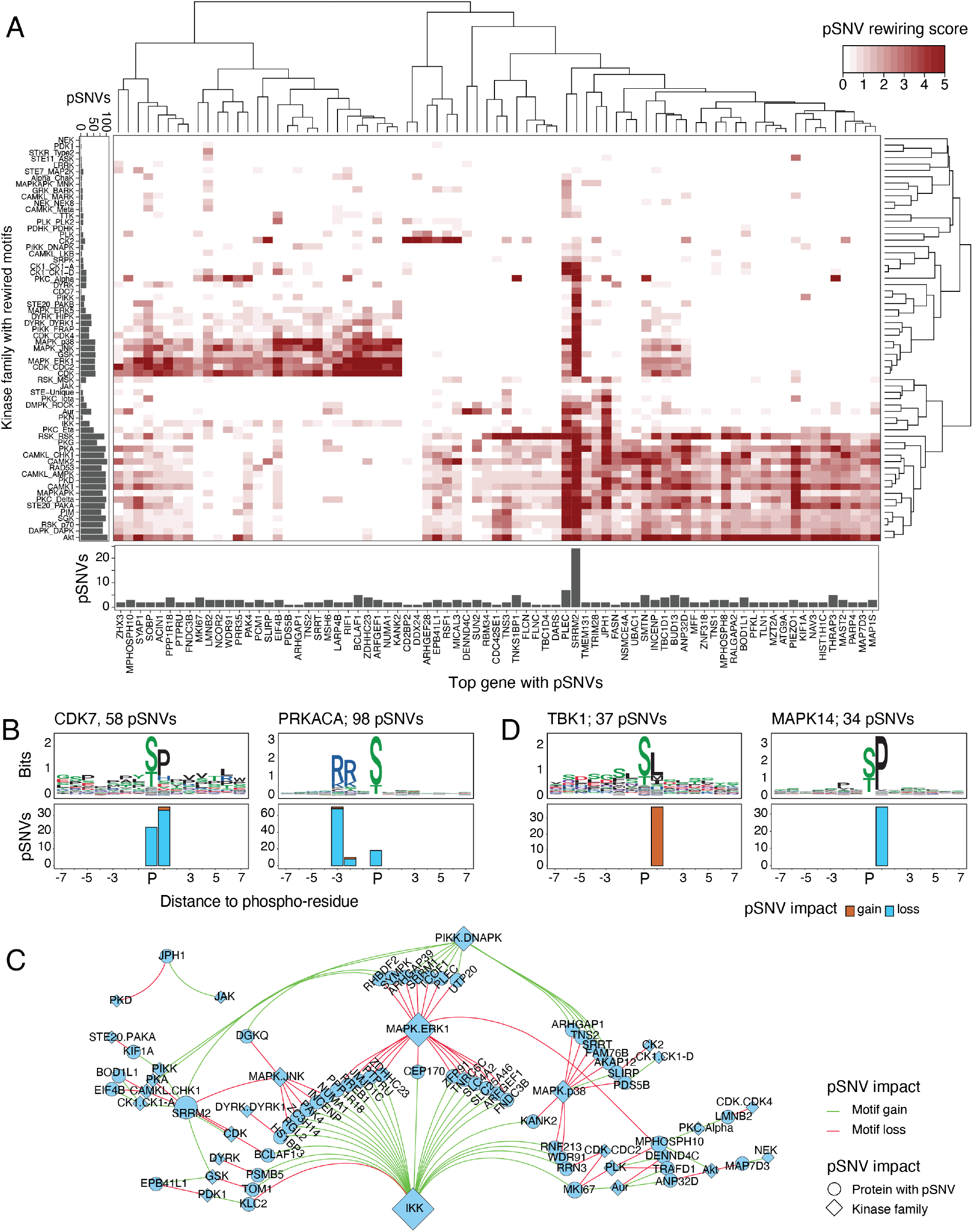
Kinases affected by pSNV-driven rewiring of sequence motifs in SARS-CoV-2 associated phosphosites. **A**. Heatmap shows the hierarchical clustering of top genes with pSNVs (X-axis), and kinase families affected by sequence motif rewiring by pSNVs (Y-axis). Genes and kinases are clustered by rewiring scores that reflect the pSNVs and their posterior probabilities of motif rewiring or central residue removal. Two clusters of kinase families are subject to motif rewiring: MAPK and CDK (middle-left) and PRKA, CAMK, CHEK, AKT, and RPS (bottom-right). Bar plots show pSNVs in sequence motifs (left) and top genes (bottom). Since motifs of different kinase families are often similar and accounted for, pSNV counts per kinase family are redundant. Scores are capped at 5 for visualization. **B**. Examples of sequence motifs rewired by pSNVs for the Cyclin Dependent Kinase 7 (CDK7) (left), Protein Kinase CAMP-Activated Catalytic Subunit Alpha (PRKACA) (middle), Calcium/Calmodulin Dependent Protein Kinase II Alpha (CAMK2A) (right). Sequence logo of the position weight matrix (PWM) bound by the kinase (top) and bar plot with pSNVs altering the motif (bottom) are shown. **C**. Network of pSNVs causing motif switches (*i*.*e*., combined gain of one motif and loss of another motif). Nodes show kinase families (diamonds) and top genes (circles), and edges show pSNV-driven motif losses and gains (red and green, respectively). Node size indicates the number of gained and lost motifs (*i*.*e*., node degree). For each pSNV, only the kinase family with the highest motif rewiring probability is shown. **D**. Sequence motifs bound by the TANK binding kinase 1 (TBK1) of the IKK family are consistently gained through pSNVs (left) while motifs bound by CDK and MAPK kinase families are lost through the same pSNVs. Bar plots show the pSNVs causing gains of TBK1 motifs (left) with corresponding losses of MAPK14 motifs (right). Similar motifs of other kinase families replaced by TBK1 motifs are not shown. pSNVs in top genes are included in panels A and B, and pSNVs in all genes are included in panels C and D.

We focused on pSNVs that disrupted one type of sequence motif and induced another type of motif at the same mutated phosphosite, potentially causing switches in kinase binding. This revealed a network of 54 proteins with pSNV-induced binding switches involving 26 kinase families, when selecting the most confident kinase family for every motif-rewiring prediction (**Figure 4C, Supplementary Table 6**). Motif switches were caused by 60 pSNVs. Strikingly, the largest subnetwork of motif switches involved motif gains of the Tank Binding Kinase 1 (TBK1) of the IKB kinase (IKK) family that replaced motifs of CDKs and MAPKs (**Figure 4D**). We found 37 pSNVs with this motif-switching impact. The top proteins with motif-switching pSNVs included the transcriptional regulators BCLAF1 and RREB1, the WDR91 protein involved in host cell entry of SARS-CoV-2 ^57^, the nuclear matrix protein NUMA, and the proliferation marker MKI67. TBK1 controls immune monitoring pathways and phosphorylates interferon regulatory factors IRF-3 and IRF-7 to trigger the host antiviral response that is suppressed by SARS-CoV-2 proteins ^10,11,14,15^. Thus, in some individuals and populations, motif-rewiring pSNVs induce structural changes in kinase signaling networks and cross-talk of mitogenic and immune response pathways. These network-rewiring pSNVs potentially reflect the inter-individual heterogeneity and evolutionary pressures of signaling networks responding to viral infections.

### Population genetic variation and disease associations of pSNVs

We studied the allele frequency of pSNVs to evaluate the extent of predicted changes in infection-responsive signaling networks in the 16 human populations in the gnomAD dataset (**Figure 5A**). Most pSNVs were relatively infrequent, with a median allele frequency of four per hundred thousand individuals (AF_all_ = 3.8 × 10^−5^) while the median frequency in the most representative population was an order of magnitude higher (AF_popmax_ = 2.1 × 10^−4^), suggesting the value of a deeper population-based analysis of certain pSNVs. Some pSNVs (86 or 4.2%) were relatively common and occurred at above 1% frequency in at least one population, including 15 pSNVs involved in sequence motif rewiring or loss of phospho-residues. For example, the pSNV P257L in WDR91 commonly observed in the human population (AF_all_ = 0.72) causes switches of sequence motifs at the phosphosite S257, by inducing motifs of the TBK1 kinase and disrupting motifs of CDK and MAPK kinases. *WDR91* encodes a WD repeat-containing protein that was identified in a CRISPR screen as a host regulator of endosomal entry of SARS-CoV-2 ^57^. Frequent motif-rewiring pSNVs were found in other top proteins, such as the spliceosome subunit SRRM2, the pro-apoptotic splicing and transcription factor BCLAF1, and the HIV-related nuclear membrane protein SUN2. pSNV frequency varied among populations in gnomAD (**Figure 5B**). For example, the pSNV S473L in TRIM28 replacing the phosphoresidue S473 involved in interferon regulation was exclusively found in the African population (AF_afr_ = 1.2 × 10^−4^). The adjacent pSNV R472C detected in Latino, North-Western European, and other populations (10^−5^ < AF < 10^−4^) may also contribute to altered signaling, however no motif disruptions were identified for this pSNV. As another example, the pSNV R585Q disrupting sequence motifs in the TBC1D4 phosphosite S588 involved in glucose transport was detected in Latino, South Asian and North-Western European populations (AF_sas_ = 1.3 × 10^−4^ and AF_amr_ = 2.9 × 10^−5^ and AF_nfe_nwe_ = 4.8 × 10^−5^) while additional pSNVs in TBC1D1 and TBC1D4 found in other populations may also contribute to the rewiring of SARS-CoV-2 associated signaling networks.

**Figure 5.**
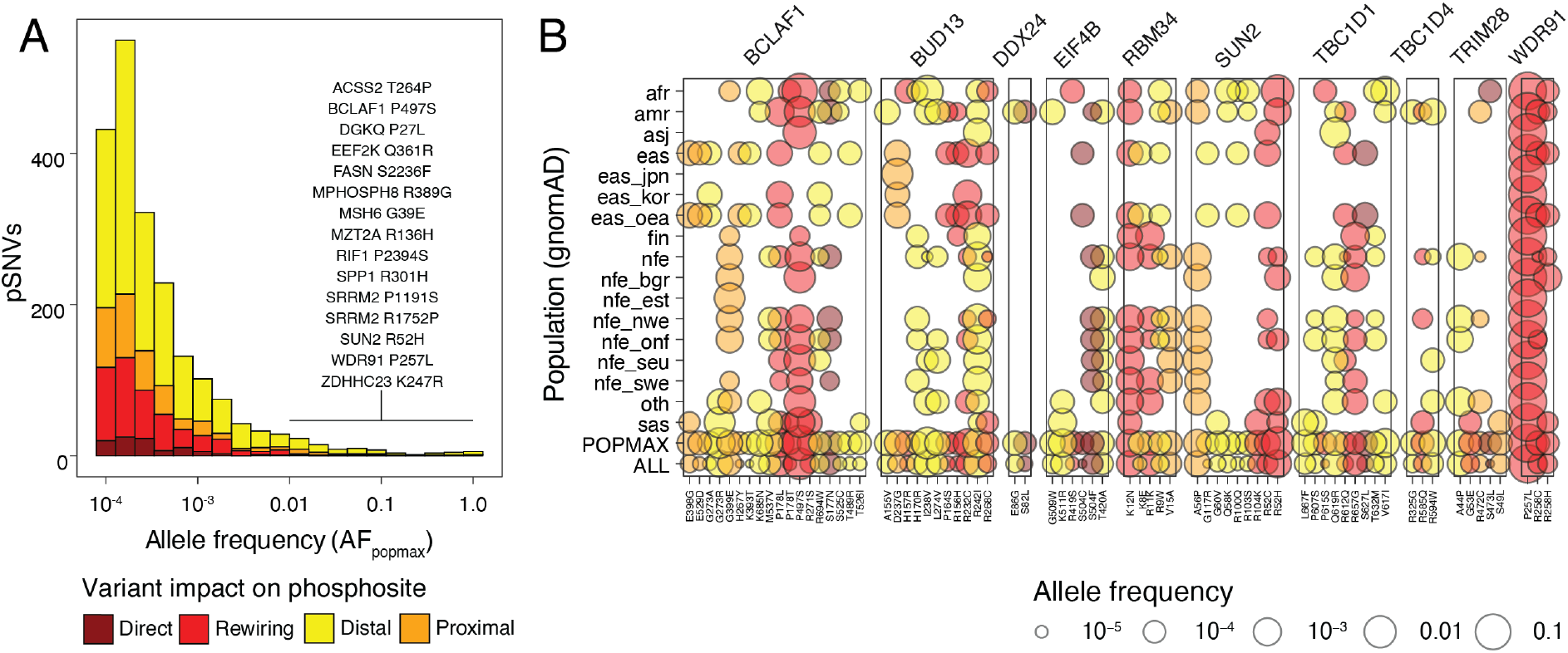
Allele frequencies of SARS-CoV-2-associated pSNVs in the human population. **A**. Histogram of allele frequencies of pSNVs in the gnomAD dataset (AF_popmax_) grouped by functional impact of pSNVs. AF_popmax_ corresponds to the population group where the allele frequency was the highest. More-frequent and impactful pSNVs are listed, with AF_popmax_ of least 1% and a direct or motif-rewiring impact. **B**. Allele frequencies of pSNVs vary among human populations in gnomAD. Dot plot shows a subset of top genes and pSNVs discussed in the study (X-axis) with their allele frequencies in populations, the maximum population-based allele frequency (AF_popmax_) and the total allele frequency in gnomAD (AF_all_) (Y-axis).

We found 86 pSNVs associated with human disease as variants of unknown significance (VUS) in the ClinVar database ^58^ (**Supplementary Table 7**). pSNV were associated with cancer predisposition (NF1, PMS2, FLCN, MSH6, APC), cardiovascular phenotypes (BMPR2, MYLK, VCL, LMNA), muscular dystrophy (SUN2, FLNC), natural killer cell and glucocorticoid deficiency (MCM4), autoimmune interstitial lung, joint, kidney disease (COPA), and renal cysts and diabetes syndrome (HNF1B). Motif-rewiring effects of disease-associated pSNVs suggest that SARS-CoV-2 and COVID-19 may hijack signal transduction networks involved in human disease. However, since the disease-associated pSNVs are annotated as variants of unknown significance, we have currently no evidence of their involvement in pathogenesis.

We asked if the pSNVs could be associated directly with risk of COVID-19, by analyzing the genome-wide association study (GWAS) of the COVID-19 Host Genetics Initiative that explored the genetic variation of nearly 50,000 COVID-19 patients using genotyping and imputation ^6^. Only a minority of pSNVs we identified were reported in the GWAS (66 or 4%). This low coverage is likely explained by the overall low allele frequency of most pSNVs that restricts their imputation and risk assessment in patient cohorts. We found a few pSNVs with nominal statistical associations with COVID-19 risk, however, the associations were not significant after multiple testing correction (all *FDR* > 0.15).

Collectively, these data suggest that pSNVs may mediate topological variation in signaling networks that respond to SARS-CoV-2 infection. Since most pSNVs occur infrequently in the human population, associating these with clinical risk using current datasets is challenging. Focused genetic association studies and mechanistic experiments are needed to confirm and characterize these predictions, and further evaluate their clinical potential.

## DISCUSSION

We highlighted human genetic polymorphisms that may cause rewiring of phospho-signaling networks responding to SARS-CoV-2 infection, using a machine-learning approach that identified impactful mutations in protein sequence motifs. The proteome-wide enrichment of motif-rewiring pSNVs suggests evolutionary plasticity of the human signaling network responding to virus infection. This evolution of host defense strategies is exemplified by the number of motif-switching pSNVs that induce novel substrate sites of the TANK Binding Kinase 1 in favor of motifs recognized by MAPK and CDK kinases. Motif-rewiring pSNVs were found in core host processes of the viral life cycle such as RNA splicing and cell cycle, innate immune responses such as interferon activation, Ras/GTPase signaling proteins, and in other human genes involved in virus infections. Genes with pSNVs were often expressed in diverse human tissues, reflecting the broad organotropism of SARS-CoV-2 and potential contribution of pSNVs to multi-organ failure in severe COVID-19, as well as diverse long-term symptoms. Disease genes and variants of unknown significance found among pSNVs suggest that infections may hijack disease pathways, and the pSNVs in the pathways may contribute to symptomatic diversity and comorbidities of COVID-19.

Our study has important limitations. We currently have no statistical or clinical evidence of pSNVs associating with SARS-CoV-2 infection, COVID-19 risk or comorbidities, as most pSNVs are infrequent in human populations and remain unmapped in genome-wide association studies. The phosphosites we studied reflect an early post-infection timepoint in cell culture, limiting our analysis of signaling pathways activated further downstream of SARS-CoV-2 infection in human tissues and the immune system. On the one hand, our analysis may overestimate the extent of network rewiring because our sequence-based pSNV impact analysis does not account for co-expression and localization of kinases and substrates. On the other hand, the number of motif-rewiring pSNVs may be underestimated, because many motifs bound by kinases and other phospho-enzymes remain unknown. The known landscape of functional protein-coding SNVs may be extended by analyzing other types of PTMs.

Our integrative proteogenomic analysis of pSNVs affecting signaling networks of SARS-CoV-2 enables further studies of viral infection, disease mechanisms and potential translation. Analysis of whole-genome sequencing datasets with matched clinical profiles of COVID-19 information is needed to associate our functional predictions of infrequent pSNVs with patient risk and co-morbidities, and to enable the development of prognostic and predictive biomarkers. The candidate genes, pathways, and kinases identified in this study enable functional genomics screens and phenotypic experiments that may lead to further mechanistic understanding of SARS-CoV-2 infection and COVID-19. The biological processes and kinase signaling networks we identified involve known drug targets, some of which have already been proposed in recent studies. Thus, our findings have the potential to contribute to genome-guided risk predictions and development of therapies for the current and future pandemics.

## METHODS

### Host phosphorylation sites

Protein phosphosites significantly differentially phosphorylated in SARS-CoV-2 infection were retrieved from the phospho-proteomics study by Bouhaddou *et al* ^9^. We mapped the phosphosites to canonical protein isoforms (hg19) using the ActiveDriverDB ^44^ database. SARS-CoV-2 associated phosphosites detected at the 24h timepoint were filtered for significance (*FDR* < 0.05 in infected cells; *FDR* > 0.05 in controls). Sites corresponding to non-phosphorylated residues in human proteins were removed (*i*.*e*., residues other than S, T, or Y). To compare observed and expected numbers of pSNVs, all experimentally detected human phosphosites were retrieved from ActiveDriverDB based on proteomics databases (PhosphoSitePlus ^59^, UniProt ^60^, Phospho.ELM ^61^, HPRD ^62^). SARS-CoV-2 associated phosphosites were excluded from these controls. Known interactions of kinases and specific phosphosites were also retrieved from ActiveDriverDB, based on multiple databases ^59-62^.

### Human genome variation

Genome variation maps of human populations were retrieved from the gnomAD exome sequencing project of 124,748 individuals (version 2.1.1; hg19) ^23^. Single nucleotide variants (SNVs) were first filtered by variant quality (*i*.*e*., by selecting variants where filter = PASS) and maximum allele frequency in the gnomAD populations (AF_popmax_ ≥ 10^−4^). The ANNOVAR software ^63^ was used to annotate the SNVs in protein-coding genes. Missense SNVs in canonical protein isoforms were selected to match the protein phosphosites. We excluded small insertions-deletions (indels), splicing, frameshift, and stop codon mutations, and missense SNVs with mismatching reference amino acid residues.

### pSNV impact prediction

Phosphorylation associated SNVs (pSNVs) occurred in SARS-CoV-2-associated phosphosites in flanking sequences of ±7 residues. Four mutually exclusive categories of variant impact were assigned in order of priority: direct (substitution of phosphorylated residues S, T, Y), motif-rewiring (gain or loss of motif, based on MIMP ^24^ analysis), and proximal and distal (within 1-2 and 3-7 residues from phosphosite, respectively, with no motif-rewiring prediction). In the cases where multiple adjacent phosphosites were found to match a pSNV, the highest-impact phosphosite was used. Motif-rewiring predictions were derived using the MIMP method ^24^, with the database of high-confidence kinase position weight matrices (PWMs) (by setting the parameter model.data = hconf), a posterior probability cutoff (*prob* > 0.5), and inclusion of central residues for rewiring predictions (include.cent = TRUE). To score each pSNV, the kinase motif (*i*.*e*., the PWM) with the largest posterior probability was selected. To prioritize pSNVs causing motif switches, both posterior probabilities (*i*.*e*., the top motif gain and loss) were selected and aggregated as *prob*_switch_ = 1 – (1 – *prob*_gain_)(1 – *prob*_loss_), corresponding to the combined probability of the pSNV causing either a gain or a loss of a motif.

### Evaluating the enrichment of motif-rewiring pSNVs

Randomly sampled phosphosites were used to evaluate the expected numbers of pSNVs with functional impact predictions (*i*.*e*., direct, motif-rewiring, proximal, and distal pSNVs). We randomly sampled 10,000 sets of phosphosites from the human phospho-proteome using datasets from ActiveDriverDB as controls, in equal numbers to SARS-CoV-2 associated phosphosites. Control sites were annotated using gnomAD SNVs and MIMP analysis, similarly to SARS-CoV-2-associated phosphosites, to derive the expected distributions of annotated pSNVs and empirical *P*-values comparing observed and expected values were reported. Expected counts were shown as mean values with ±1 standard deviation for confidence intervals.

### Gene prioritization

Genes were scored based on the aggregated impact of pSNVs in phosphosites. Each pSNV was assigned either the posterior probability of motif rewiring (*prob*_SNV_ = *prob*_rewiring_ from MIMP; see above), or alternatively, a conservative posterior probability of motif rewiring (*prob*_SNV_ = 0.05) if it was annotated as distal or proximal to the phosphosite with no motif-rewiring predictions from MIMP. To derive gene-level significance scores (*P*-values), posterior probabilities of all pSNVs in each gene were aggregated as *P*_gene_ = 1 – Π _SNV of gene_ (1-*prob*_SNV_), reflecting the null hypothesis that none of the pSNVs in the protein encoded by the gene independently altered kinase binding motifs. The resulting gene-level *P*-values were corrected for multiple testing with the Benjamini-Hochberg false discovery rate (*FDR*) method and genes with significant *FDR* values were selected (*FDR* < 0.1). *FDR* values were capped at 10^−16^ for visualization.

### Functional enrichment analysis

Pathway enrichment analysis was conducted using the ActivePathways method ^37^ and included gene sets corresponding to biological processes of Gene Ontology ^64^, molecular pathways of Reactome ^65^, protein complexes of CORUM ^66^, and genes expressed in human tissues of the Human Protein Atlas ^39^. Gene sets were retrieved from the g:Profiler web server ^67^ (Mar 25^th^, 2021). Tissue-specific gene expression signatures and functional gene sets were analyzed separately. As the statistical background, only experimentally determined human phosphoproteins in ActiveDriverDB were used for increased stringency. Significantly enriched pathways were detected using default parameters of ActivePathways and significant gene sets were selected (ActivePathways *FDR* < 0.1). Functional gene sets were visualized as an enrichment map ^68^, functional themes were annotated manually, and the highest-ranking genes with pSNVs were shown (gene *FDR* < 0.25). Genes with pSNVs highly expressed in human tissues were visualized separately. *FDR* values were capped at 10^−16^ for visualization.

### Analysis of protein-protein interaction (PPI) networks

Three types of PPIs were included to analyze the network of top genes with pSNVs (*FDR* < 0.1). First, we included physical human-human PPIs where both interacting host proteins were part of the top gene list. Second, we included physical human-virus PPIs where one interacting host protein was part of the top gene list, and the other protein was encoded by the SARS-CoV-2 virus. Third, we included experimentally verified site-specific kinase-substrate interactions where the substrate host protein was part of the top gene list, the host kinase protein was verified to bind a phosphosite of the substrate protein based on previous experimental studies, and the phosphosite had at least one pSNV. Human-human and human-virus PPIs were retrieved from the BioGRID database ^43^ (V 4.4.199, downloaded on June 29^th^, 2021). PPIs of the third type were retrieved from ActiveDriverDB. The PPI network was visualized using the Cytoscape software ^69^. Empirical *P*-values were used to evaluate the enrichment of PPIs among the top genes with pSNVs. Control sets of proteins were randomly drawn from the human phospho-proteome in equal numbers to top genes with pSNVs and control PPI networks were constructed. PPI counts in these networks provided the expected distributions and empirical *P*-values for human-human PPIs, human-virus PPIs, and human kinase-substrate PPIs with pSNVs. Expected counts were shown as mean values with ±1 standard deviation for confidence intervals.

### Analysis of population frequency and disease annotations of pSNVs

Allele frequencies of pSNVs in human populations were reported as shown in the gnomAD dataset. We also studied disease associations of pSNVs recorded in the ClinVar database ^58^ that were previously mapped to canonical protein isoforms in ActiveDriverDB. Disease annotations of pSNVs were then reviewed manually using the ClinVar website (data retrieved on Oct 6^th^, 2021). Based on this review, lower-confidence pSNV annotations with no ClinVar star ratings, and annotations with consensus rating of *benign* were filtered.

## Data Availability

All data produced in the present study are available in supplementary materials or upon reasonable request to the authors.

## SUPPLEMENTARY INFORMATION

**Supplementary Table 1. Phosphorylation associated SNVs (pSNVs) of SARS-CoV-2 infection**. The table shows all the pSNVs in the phosphosites affected in SARS-CoV-2 infection in the gnomAD dataset detected in at least one human population (AF_popmax_ ≥ 10^−4^). The table includes allele frequencies of pSNVs in the 16 populations in gnomAD (columns *AF_all, AF_popmax, etc*.), the kinases known to target the site (column *ptm_kinase*), the probability of the pSNV disrupting the phosphosite based on MIMP analysis (column *top_MIMP_score;* the highest-ranking kinase motif), the predicted impact of the pSNV on the phosphorylation (column *pSNV_impact*), and whether the pSNV affects multiple phosphosites (column *multi_PTM*). In case of multiple sites, the phosphosite with highest MIMP probability is used for scoring (see Supplementary Table 2).

**Supplementary Table 2. Motif-rewiring predictions of pSNVs**. Predictions of the impact of pSNVs on sequence motifs bound by kinases, derived from the MIMP method. Provided are the amino acids in the phosphosites of reference and mutated sequences, the corresponding MIMP scores of the reference and mutated sequences of being bound by kinases, the ratio of scores (column *log_ratio*), and the posterior probability of motif rewiring (column *prob*). Missing values (NA) are shown for pSNV scores where the impact is direct removal of a phosphoresidue.

**Supplementary Table 3. Top genes with pSNVs**. Complete list of genes with pSNVs ranked by their *FDR* scores, corresponding to the null hypothesis that no pSNVs per gene cause a motif rewiring at SARS-CoV-2-associated phosphosites. Filtered genes (*FDR* < 0.1) are used in most analyses.

**Supplementary Table 4. Molecular pathways, processes, and tissue-specific gene expression profiles of top genes with pSNVs**. Results of pathway enrichment analysis of top genes with pSNV using ActivePathways (*FDR* < 0.1). Gene sets of biological processes of Gene Ontology, molecular pathways of Reactome, protein complexes of CORUM, and tissue specific gene expression signatures from the Human Protein Atlas (HPA) database are shown. For each enriched gene set, the *FDR*-ranked list of genes with pSNVs in the gene set is included.

**Supplementary Table 5. Protein-protein interaction (PPI) network of top genes with pSNVs**. Three types of interactions are shown (column *interaction_type*): physical interactions of top genes (proteins) with pSNVs (*i*.*e*., *human_human*), physical interactions of top genes with pSNVs and proteins encoded by SARS-CoV-2 (*i*.*e*., *human_sars*), and site-specific signaling interactions of top genes with pSNVs and known kinases binding these phosphosites based on information collected in ActiveDriverDB (*i*.*e*., *w_kinase*). Site-specific interactions have at least one pSNV in the phosphosites. The scores are -log10-transformed *FDR* values of top genes.

**Supplementary Table 6. Motif-switching pSNVs in top genes**. The subset of pSNVs causing motif switching, resulting in the joint loss of one type of motif and the gain of another motif. For each pSNV, only the kinase and kinase family of the highest-scoring prediction is shown (see Supplementary Table 2 for all predictions).

**Supplementary Table 7. Disease annotations of all pSNVs**. Disease annotations of pSNVs from the ClinVar database are shown. Consensus evaluation of disease significance was based on a systematic analysis of ClinVar annotations in ActiveDriverDB and a manual review of the ClinVar website (column *significance_curated*). Most disease-associated pSNVs are annotated as variants of unknown significance (VUS). Multiple disease annotations of the same pSNV are concatenated (column *disease*).

## Acknowledgments

This work was supported by the COVID-19 Supplements of the Project Grant of Canadian Institutes of Health Research (CIHR) to J.R. and the Discovery Grant of the Natural Sciences and Engineering Research Council (NSERC) to J.R., as well as the Investigator Award to J.R. from the Ontario Institute for Cancer Research (OICR). A.B. was supported by an Ontario Graduate Scholarship. M.K. was supported by the Scatcherd European Scholarship. Funding to OICR is provided by the Government of Ontario.

## Author contributions

D.P., A.B. and J.R. analyzed the data, interpreted the results, and prepared the figures. M.K. preprocessed the data and led the database development. D.P., A.B., and J.R. wrote the manuscript. J.R. conceived and supervised the project. All authors reviewed and edited the manuscript and approved the final version.

## Notes

### Competing Interest Statement

The authors have declared no competing interest.

### Author Declarations

This study involves only openly available human data, which can be obtained from the gnomAD database.

## References

1 Richardson, S. et al. Presenting Characteristics, Comorbidities, and Outcomes Among 5700 Patients Hospitalized With COVID-19 in the New York City Area. JAMA 323, 2052–2059, doi:10.1001/jama.2020.6775 (2020).

2 Webb Hooper, M., Napoles, A. M. & Perez-Stable, E. J. COVID-19 and Racial/Ethnic Disparities. JAMA 323, 2466–2467, doi:10.1001/jama.2020.8598 (2020).

3 Pareek, M. et al. Ethnicity and COVID-19: an urgent public health research priority. The Lancet 395, 1421–1422, doi:10.1016/s0140-6736(20)30922-3 (2020).

4 Nicola, M. et al. The socio-economic implications of the coronavirus pandemic (COVID-19): A review. Int J Surg 78, 185–193, doi:10.1016/j.ijsu.2020.04.018 (2020).

5 Severe Covid Gwas Group et al. Genomewide Association Study of Severe Covid-19 with Respiratory Failure. N Engl J Med 383, 1522–1534, doi:10.1056/NEJMoa2020283 (2020).

6 Covid-19 Host Genetics Initiative. Mapping the human genetic architecture of COVID-19. Nature, doi:10.1038/s41586-021-03767-x (2021).

7 Pairo-Castineira, E. et al. Genetic mechanisms of critical illness in COVID-19. Nature 591, 92–98, doi:10.1038/s41586-020-03065-y (2021).

8 Vidal, M., Cusick, M. E. & Barabasi, A. L. Interactome networks and human disease. Cell 144, 986–998, doi:10.1016/j.cell.2011.02.016 (2011).

9 Bouhaddou, M. et al. The Global Phosphorylation Landscape of SARS-CoV-2 Infection. Cell 182, 685–712 e619, doi:10.1016/j.cell.2020.06.034 (2020).

10 Sharma, S. et al. Triggering the interferon antiviral response through an IKK-related pathway. Science 300, 1148–1151, doi:10.1126/science.1081315 (2003).

11 Balka, K. R. et al. TBK1 and IKKepsilon Act Redundantly to Mediate STING-Induced NF-kappaB Responses in Myeloid Cells. Cell Rep 31, 107492, doi:10.1016/j.celrep.2020.03.056 (2020).

12 Matsuyama, T., Kubli, S. P., Yoshinaga, S. K., Pfeffer, K. & Mak, T. W. An aberrant STAT pathway is central to COVID-19. Cell Death Differ 27, 3209–3225, doi:10.1038/s41418-020-00633-7 (2020).

13 Appelberg, S. et al. Dysregulation in Akt/mTOR/HIF-1 signaling identified by proteo-transcriptomics of SARS-CoV-2 infected cells. Emerg Microbes Infect 9, 1748–1760, doi:10.1080/22221751.2020.1799723 (2020).

14 Lei, X. et al. Activation and evasion of type I interferon responses by SARS-CoV-2. Nat Commun 11, 3810, doi:10.1038/s41467-020-17665-9 (2020).

15 Xia, H. et al. Evasion of Type I Interferon by SARS-CoV-2. Cell Rep 33, 108234, doi:10.1016/j.celrep.2020.108234 (2020).

16 Hirano, T. & Murakami, M. COVID-19: A New Virus, but a Familiar Receptor and Cytokine Release Syndrome. Immunity 52, 731–733, doi:10.1016/j.immuni.2020.04.003 (2020).

17 Mishra, S., Bassi, G. & Nyomba, B. G. Inter-proteomic posttranslational modifications of the SARS-CoV-2 and the host proteins A new frontier. Exp Biol Med (Maywood) 246, 749–757, doi:10.1177/1535370220986785 (2021).

18 Li, S., Iakoucheva, L. M., Mooney, S. D. & Radivojac, P. Loss of post-translational modification sites in disease. Pac Symp Biocomput, 337–347 (2010).

19 Reimand, J. & Bader, G. D. Systematic analysis of somatic mutations in phosphorylation signaling predicts novel cancer drivers. Molecular systems biology 9, 637, doi:10.1038/msb.2012.68 (2013).

20 Creixell, P. et al. Kinome-wide decoding of network-attacking mutations rewiring cancer signaling. Cell 163, 202–217, doi:10.1016/j.cell.2015.08.056 (2015).

21 Huang, K. L. et al. Pathogenic Germline Variants in 10,389 Adult Cancers. Cell 173, 355–370 e314, doi:10.1016/j.cell.2018.03.039 (2018).

22 Reimand, J., Wagih, O. & Bader, G. D. Evolutionary constraint and disease associations of post-translational modification sites in human genomes. PLoS Genet 11, e1004919, doi:10.1371/journal.pgen.1004919 (2015).

23 Karczewski, K. J. et al. The mutational constraint spectrum quantified from variation in 141,456 humans. Nature 581, 434–443, doi:10.1038/s41586-020-2308-7 (2020).

24 Wagih, O., Reimand, J. & Bader, G. D. MIMP: predicting the impact of mutations on kinase-substrate phosphorylation. Nat Methods 12, 531–533, doi:10.1038/nmeth.3396 (2015).

25 Wojcechowskyj, J. A. et al. Quantitative phosphoproteomics reveals extensive cellular reprogramming during HIV-1 entry. Cell Host Microbe 13, 613–623, doi:10.1016/j.chom.2013.04.011 (2013).

26 Zhang, X. et al. Structure of the human activated spliceosome in three conformational states. Cell Res 28, 307–322, doi:10.1038/cr.2018.14 (2018).

27 Frankiw, L. et al. BUD13 Promotes a Type I Interferon Response by Countering Intron Retention in Irf7. Mol Cell 73, 803–814 e806, doi:10.1016/j.molcel.2018.11.038 (2019).

28 Banerjee, A. K. et al. SARS-CoV-2 Disrupts Splicing, Translation, and Protein Trafficking to Suppress Host Defenses. Cell 183, 1325–1339 e1321, doi:10.1016/j.cell.2020.10.004 (2020).

29 Finkel, Y. et al. SARS-CoV-2 uses a multipronged strategy to impede host protein synthesis. Nature 594, 240–245, doi:10.1038/s41586-021-03610-3 (2021).

30 Ostaszewski, M. et al. COVID19 Disease Map, a computational knowledge repository of virus-host interaction mechanisms. Mol Syst Biol 17, e10387, doi:10.15252/msb.202110387 (2021).

31 Qin, C. et al. Bclaf1 critically regulates the type I interferon response and is degraded by alphaherpesvirus US3. PLoS Pathog 15, e1007559, doi:10.1371/journal.ppat.1007559 (2019).

32 McPherson, J. P. et al. Essential role for Bclaf1 in lung development and immune system function. Cell Death Differ 16, 331–339, doi:10.1038/cdd.2008.167 (2009).

33 Krischuns, T. et al. Phosphorylation of TRIM28 Enhances the Expression of IFN-beta and Proinflammatory Cytokines During HPAIV Infection of Human Lung Epithelial Cells. Front Immunol 9, 2229, doi:10.3389/fimmu.2018.02229 (2018).

34 Hu, C. et al. Roles of Kruppel-associated Box (KRAB)-associated Co-repressor KAP1 Ser-473 Phosphorylation in DNA Damage Response. The Journal of biological chemistry 287, 18937–18952, doi:10.1074/jbc.M111.313262 (2012).

35 Wang, Y. et al. TRIM28 regulates SARS-CoV-2 cell entry by targeting ACE2. Cell Signal 85, 110064, doi:10.1016/j.cellsig.2021.110064 (2021).

36 Tovo, P. A. et al. COVID-19 in Children: Expressions of Type I/II/III Interferons, TRIM28, SETDB1, and Endogenous Retroviruses in Mild and Severe Cases. Int J Mol Sci 22, doi:10.3390/ijms22147481 (2021).

37 Paczkowska, M. et al. Integrative pathway enrichment analysis of multivariate omics data. Nature Communications 11, 735 (2020).

38 Knoops, K. et al. SARS-coronavirus replication is supported by a reticulovesicular network of modified endoplasmic reticulum. PLoS Biol 6, e226, doi:10.1371/journal.pbio.0060226 (2008).

39 Uhlen, M. et al. Proteomics. Tissue-based map of the human proteome. Science 347, 1260419, doi:10.1126/science.1260419 (2015).

40 Feng, Z. et al. The Novel Severe Acute Respiratory Syndrome Coronavirus 2 (SARS-CoV-2) Directly Decimates Human Spleens and Lymph Nodes. doi:10.1101/2020.03.27.20045427 (2020).

41 Yang, A. C. et al. Dysregulation of brain and choroid plexus cell types in severe COVID-19. Nature 595, 565–571, doi:10.1038/s41586-021-03710-0 (2021).

42 Gupta, A. et al. Extrapulmonary manifestations of COVID-19. Nat Med 26, 1017–1032, doi:10.1038/s41591-020-0968-3 (2020).

43 Oughtred, R. et al. The BioGRID interaction database: 2019 update. Nucleic Acids Res 47, D529–D541, doi:10.1093/nar/gky1079 (2019).

44 Krassowski, M. et al. ActiveDriverDB: Interpreting Genetic Variation in Human and Cancer Genomes Using Post-translational Modification Sites and Signaling Networks (2021 Update). Front Cell Dev Biol 9, 626821, doi:10.3389/fcell.2021.626821 (2021).

45 Popkin, B. M. et al. Individuals with obesity and COVID-19: A global perspective on the epidemiology and biological relationships. Obes Rev 21, e13128, doi:10.1111/obr.13128 (2020).

46 Baruah, C., Devi, P. & Sharma, D. K. Sequence Analysis and Structure Prediction of SARS-CoV-2 Accessory Proteins 9b and ORF14: Evolutionary Analysis Indicates Close Relatedness to Bat Coronavirus. Biomed Res Int 2020, 7234961, doi:10.1155/2020/7234961 (2020).

47 Ma, J. et al. The requirement of the DEAD-box protein DDX24 for the packaging of human immunodeficiency virus type 1 RNA. Virology 375, 253–264, doi:10.1016/j.virol.2008.01.025 (2008).

48 Almasy, K. M., Davies, J. P. & Plate, L. Comparative host interactomes of the SARS-CoV-2 nonstructural protein 3 and human coronavirus homologs. bioRxiv, doi:10.1101/2021.03.08.434440 (2021).

49 Sakai, Y. et al. Two-amino acids change in the nsp4 of SARS coronavirus abolishes viral replication. Virology 510, 165–174, doi:10.1016/j.virol.2017.07.019 (2017).

50 Cottam, E. M. et al. Coronavirus nsp6 proteins generate autophagosomes from the endoplasmic reticulum via an omegasome intermediate. Autophagy 7, 1335–1347, doi:10.4161/auto.7.11.16642 (2011).

51 Sano, H. et al. Insulin-stimulated phosphorylation of a Rab GTPase-activating protein regulates GLUT4 translocation. The Journal of biological chemistry 278, 14599–14602, doi:10.1074/jbc.C300063200 (2003).

52 Stone, S. et al. TBC1D1 is a candidate for a severe obesity gene and evidence for a gene/gene interaction in obesity predisposition. Hum Mol Genet 15, 2709–2720, doi:10.1093/hmg/ddl204 (2006).

53 Moltke, I. et al. A common Greenlandic TBC1D4 variant confers muscle insulin resistance and type 2 diabetes. Nature 512, 190–193, doi:10.1038/nature13425 (2014).

54 Coffey, S., Costacou, T., Orchard, T. & Erkan, E. Akt Links Insulin Signaling to Albumin Endocytosis in Proximal Tubule Epithelial Cells. PLoS One 10, e0140417, doi:10.1371/journal.pone.0140417 (2015).

55 Geraghty, K. M. et al. Regulation of multisite phosphorylation and 14-3-3 binding of AS160 in response to IGF-1, EGF, PMA and AICAR. Biochem J 407, 231–241, doi:10.1042/BJ20070649 (2007).

56 Blasius, M. et al. A phospho-proteomic screen identifies substrates of the checkpoint kinase Chk1. Genome Biol 12, R78, doi:10.1186/gb-2011-12-8-r78 (2011).

57 Zhu, Y. et al. A genome-wide CRISPR screen identifies host factors that regulate SARS-CoV-2 entry. Nat Commun 12, 961, doi:10.1038/s41467-021-21213-4 (2021).

58 Landrum, M. J. et al. ClinVar: improvements to accessing data. Nucleic Acids Res 48, D835–D844, doi:10.1093/nar/gkz972 (2020).

59 Hornbeck, P. V. et al. PhosphoSitePlus, 2014: mutations, PTMs and recalibrations. Nucleic Acids Res 43, D512–520, doi:10.1093/nar/gku1267 (2015).

60 UniProt Consortium. UniProt: a worldwide hub of protein knowledge. Nucleic Acids Res 47, D506–D515, doi:10.1093/nar/gky1049 (2019).

61 Dinkel, H. et al. Phospho.ELM: a database of phosphorylation sites--update 2011. Nucleic acids research 39, D261–267, doi:10.1093/nar/gkq1104 (2011).

62 Keshava Prasad, T. S. et al. Human Protein Reference Database--2009 update. Nucleic acids research 37, D767–772, doi:10.1093/nar/gkn892 (2009).

63 Wang, K., Li, M. & Hakonarson, H. ANNOVAR: functional annotation of genetic variants from high-throughput sequencing data. Nucleic acids research 38, e164, doi:10.1093/nar/gkq603 (2010).

64 Ashburner, M. et al. Gene ontology: tool for the unification of biology. The Gene Ontology Consortium. Nature genetics 25, 25–29, doi:10.1038/75556 (2000).

65 Fabregat, A. et al. The Reactome Pathway Knowledgebase. Nucleic Acids Res 46, D649–D655, doi:10.1093/nar/gkx1132 (2018).

66 Ruepp, A. et al. CORUM: the comprehensive resource of mammalian protein complexes--2009. Nucleic acids research 38, D497–501, doi:10.1093/nar/gkp914 (2010).

67 Reimand, J., Kull, M., Peterson, H., Hansen, J. & Vilo, J. g:Profiler--a web-based toolset for functional profiling of gene lists from large-scale experiments. Nucleic acids research 35, W193–200, doi:10.1093/nar/gkm226 (2007).

68 Reimand, J. et al. Pathway enrichment analysis and visualization of omics data using g:Profiler, GSEA, Cytoscape and EnrichmentMap. Nat Protoc 14, 482–517, doi:10.1038/s41596-018-0103-9 (2019).

69 Cline, M. S. et al. Integration of biological networks and gene expression data using Cytoscape. Nature protocols 2, 2366–2382, doi:10.1038/nprot.2007.324 (2007).

